# Analysis on Action Tracking Reports of COVID-19 Informs Control Strategies and Vaccine Delivery in Post-Pandemic Era

**DOI:** 10.1101/2021.04.08.21254953

**Authors:** Xiaofei Sun, Tianjia Guan, Tao Xue, Chun Fan, Meng Yang, Yuxian Meng, Tianwei Zhang, Bahabaike Jiangtulu, Fei Wu, Jiwei Li

## Abstract

Understanding the spread of SARS-CoV-2 provides important insights for control policies such as social-distancing interventions and vaccine delivery in the post-pandemic era. In this work, we take the advantage of action tracking reports of confirmed COVID-19 patients, which contain the mobility trajectory of patients. We analyzed reports of patients from April 2020 to January 2021 in China, a country where the residents are well-prepared for the “new normal” world following COVID-19 spread. We developed natural language processing (NLP) tools to transform the unstructured text of action-tracking reports to a structured network of social contacts. An epidemiology model was built on top of the network. Our analysis provides important insights for the development of control policies. Under the “new normal” conditions, we find that restaurants, locations less protected by mask-wearing, have a greater risk than any other location categories, including locations where people are present at higher densities (e.g., flight). We find that discouraging railway transports is crucial to avoid another wave of breakout during the Chunyun season (a period of travel in China with extremely high traffic load around the Chinese New Year). By formalizing the challenge of finding the optimal vaccine delivery among various different population groups as an optimization problem, our analysis helps to maximize the efficiency of vaccine delivery under the general situation of vaccine supply shortage. We are able to reduce the numbers of infections and deaths by 7.4% and 10.5% respectively with vaccine supply for only 1% of the population. Furthermore, with 10% vaccination rate, the numbers of infections and deaths further decrease by 52.6% and 78.1% respectively. Our work will be helpful in the design of effective policies regarding interventions, reopening, contact tracing and vaccine delivery in the “new normal” world following COVID-19 spread.

## 1 Main

Understanding the spread of SARS-CoV-2 is important in the “new normal” world following COVID-19 spread: first, it provides important insights regarding control policies, such as when and which relocations should be reopened, which are important for balancing disruptions caused by interventions and reducing transmission. Second, it informs more effective contact tracing strategies, regarding which groups of people should be tested immediately or quarantined if they have interactions with a confirmed case. Third, it informs strategies for effective vaccine delivery under the conditions of vaccine shortage.

Many recent efforts have been devoted to modeling SARS-CoV-2 transmission [1, 2, 3, 4, 5, 6, 7, 8, 9]. These recent efforts have two main limitations. First, there is a lack of first-hand tracking details: existing approaches are mostly focused on aggregate historical data [10, 11, 12] and simulated data [13, 14], and mobility data [15, 16, 17, 18, 19, 20, 21]. [20] proposed a method to learn the transmission model by fitting the number of city-level confirmed cases based on the mobility data. However, mobility data does not contain statistics regarding the number of infections associated with visiting certain locations. These statistics must be regarded as unknown variables to be further learned by fitting the city-level infection number, which inevitably introduces substantial noise. Therefore, the model trained based only on mobility data is incapable of capturing these important aspects; Second, existing efforts are mostly concerned with the first wave of outbreaks rather than the prolonged pandemic with altered day-to-day human behavior patterns (e.g., mask-wearing). Results and conclusions cannot be extended to the post-pandemic era, such as the situation in China after April 2020, when the pandemic was generally under control with only sporadic local outbreaks.

Analyzing actions and tracking of real patients with COVID-19 can facilitate the development of more accurate models of the spread of SARS-CoV-2, and thus inform more effective policy responses. Here, we propose analyzing action tracking reports of confirmed cases in China from Apr 2020 to Jan 2021, a period during which the residents are well-prepared for the “new normal” world following COVID-19 spread. Briefly, action tracking reports contain the mobility trajectories of patients with COVID-19 within the period of 3-14 days before diagnosis. They were published to the public to warn local residents of places that confirmed patients have been to, but are carefully phrased and structured to preserve the privacy of confirmed patients and make sure that patients are unidentifiable.^1^ Action tracking reports serve as a valuable data foundation for understanding the spread of COVID-19 to inform control and vaccine delivery strategies. We collected reports for a total number of 1,752 patients from Apr 2020 to Jan 2021 in China, representing approximately 20% of all confirmed cases within this period.

**Figure 1:**
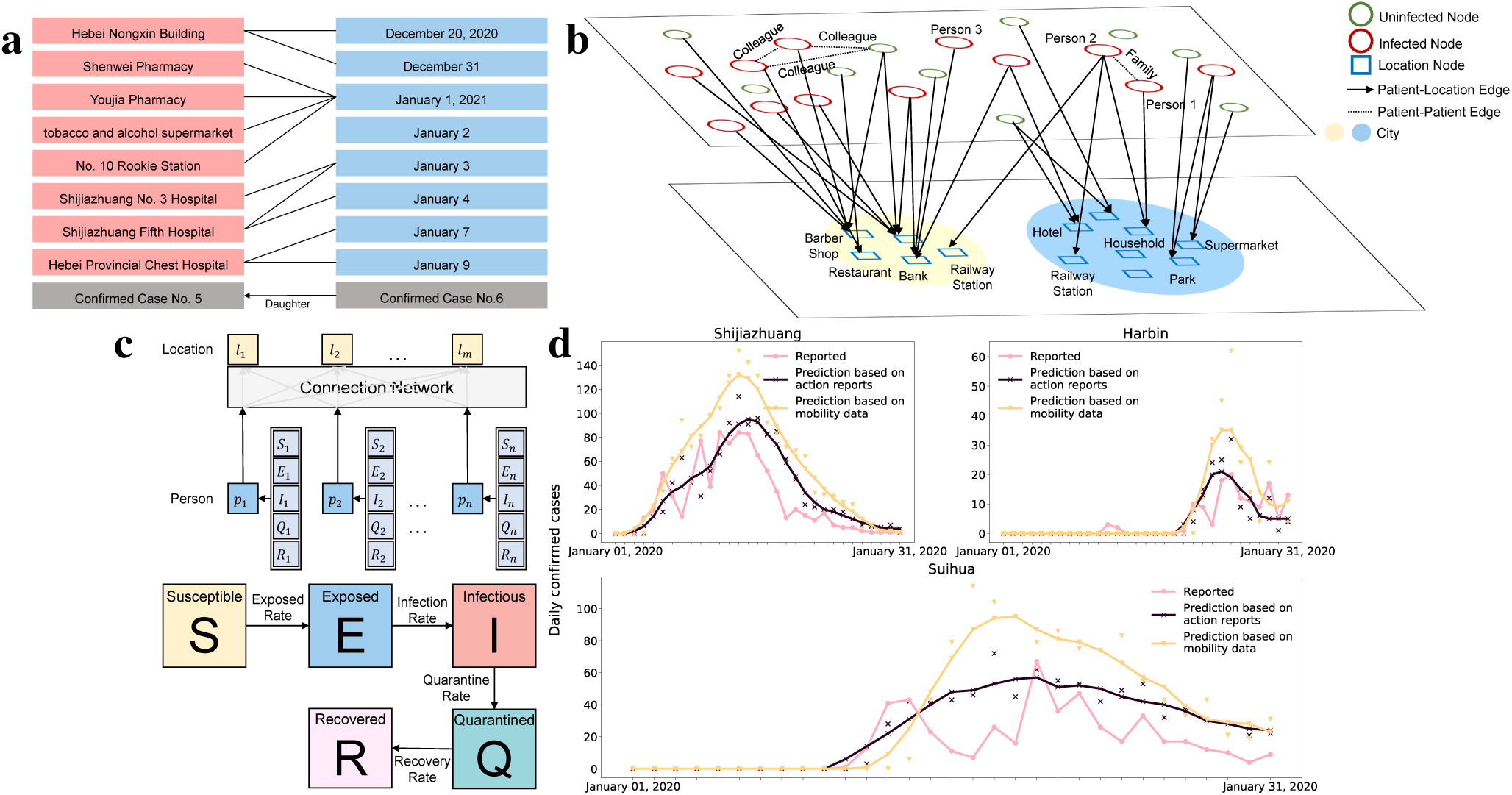
**a**, Using natural language processing (NLP) models to extract structural information from action tracking reports. Entities including location and time are first extracted from raw tracking reports. Then, based on the extracted entities, location-time relationships are constructed, forming structured edges between locations and time points. **b**, Construction of a mobility network based on the extracted structured information. A node in the network represents a patient or a location, and time-varying edges are constructed between a patient node and a location node if the patient visited that location at a given time point. The network also contains statistics of uninfected nodes collected from SmartSteps. Location nodes are divided into 11 categories. **c**, An SEIR model is trained based on the constructed network by fitting the number of patients visiting certain locations. Each category of location *C* is associated with a specific time-consistent transmission rate *β*_*C*_ in the SEIR model. **d**, Daily reported cases, predictions made by the proposed model based on action reports, and the predictions made by the baseline model trained based only on city-level reported cases and mobility data. Curves are smoothed by 5-day average. As shown in the figure, the model based on action reports can make predictions that are significantly more accurate than the baseline model.

We developed natural language processing (NLP) tools to transform the unstructured text of action tracking reports to a structured network (shown in Fig.1b), where a time-varying edge connects a person to a location if the patient went to that location at a given time. An SEIR model is built on top of the network to characterize the spread of coronavirus. Under the “new normal” condition, an important factor for transmission rates of different location categories is the strictness of mask-wearing enforcement, transmission rates are greater for restaurants than for other location categories, and traveling by air is safer than by rail. We demonstrate how demographic factors (e.g., age, sex and cities of different economic tiers in China) affect the transmission of coronavirus, and show that the third- and fourth-tier cities have higher infection rates, compared with the first-tier cities in China. Our study also informs effective strategies for vaccine delivery under conditions of vaccine shortage. By transforming the problem of finding the optimal vaccine delivery strategy among diverse population groups to an optimization problem, our study showed that with vaccine supply for only 1% of the population, we are able to decrease total infections by 7.4% and total deaths by 10.5%. Furthermore, with 10% vaccination rate, we are able to decrease total infections by 52.6% and total deaths by 78.1%.

### Network Construction Based on Action tracking Reports

Action tracking reports exist in a noisy form of large text chunks, making direct analysis difficult. To address this issue, we developed natural language processing (NLP) tools to transform the unstructured text of action tracking reports to structured networks, as shown in Fig.1b. The developed NLP models automatically extract entities of locations visited by patients, ted such as restaurants, railway stations, supermarkets, hotels, etc, along with the date of the visit. We construct networks based on extracted structured information for patients, where a node in the network represents a patient or a location (e.g., restaurant, supermarket). A location node has the attribute of its category. Time-varying edges are constructed between a patient node and a location node if the patient visited the location at time *t*. The current network contains only infected people among the visits, and we need statistics regarding other visits not associated with infections. To achieve this goal, we used SmartSteps, a service that aggregates anonymized location data to obtain the number of visits to a location within each time period. We divide extracted locations into 11 categories, eight of which are community locations, i.e., households, workplaces, hotels, shopping-centers/supermarkets, banks, restaurants, parks, and barber shops/hairdressers, The remaining three are associated with transportation: railway stations/trains, buses/taxis, and airports/airplanes.

### SEIR Model Based on Constructed Network

We built the susceptible–exposed–infectious–removed (SEIR) model on top of the constructed network (Fig.1c). Extracted from the reports, the actual quarantine period for confirmed cases is readily identified in the network. Because action tracking reports contain (almost) all locations that confirmed patients have been to, and because the number of reports constitutes a relatively large proportion of all infected cases in China, we assume that contacts between susceptible populations and exposed cases only occur in the locations mentioned in the action tracking reports of confirmed patients. ^2^ To characterize the transmission risks of different categories of locations, each category of location *C* is associated with a category specific transmission rate *β*_*C*_ in the SEIR model. *β*_*C*_ are time-consistent and free parameters to be learned based on network observations. The model is optimized using the least-square regression to minimize the gap between confirmed case counts for each location and predicted number of confirmed cases for that location. Given the trained SEIR model with learned parameters *β*_*C*_, we can examine the impacts of different policies by running the SEIR model on the counterfactual network with features corresponding to the policy.

### Risks of Different Location Categories

Here we define *R*_*c*_, which denotes the number of secondary patients caused by a patient at location *c* during this single visit if the patient visits location *c. R*_*c*_ can be estimated based on the dataset. As shown in Fig.2a, the highest value of *R*_*c*_ was obtained for restaurants, followed by supermarkets/grocery-stores, railway-stations/trains, hotels, buses/taxis, workplaces, barber shops/hairdressers, parks, banks and airports/airplanes. Restaurants and supermarkets/grocery stores have the highest reproduction numbers (R), and restaurants consistently carry greater risks than supermarkets/grocery stores. This observation is different from the findings in a recent study [20], where restaurants contributed less to infections over time because of lockdowns, but the contributions of supermarkets/grocery-stores remained steady or even increased because they are considered as essential businesses. These differences between observations are presumably because for the study period in this work, mask-wearing has been adopted by almost the entire population. This considerably differs from the situation considered in the previous research [20]. Therefore, an important factor for the virus transmission is the strictness of mask-wearing enforcement in certain types of locations. In restaurants, people tend not to wear masks while eating, and are therefore exposed to greater transmission risks than in grocery-stores. Surprisingly, the transmission rates for airports/airplanes are lower than other means of transportation (e.g., trains and buses), as well as other community locations (e.g., restaurants and hotels). We postulate that this is because mask-wearing is strictly enforced on airplanes and in airports, but less strictly on trains and buses during longer periods of transportation.

**Figure 2:**
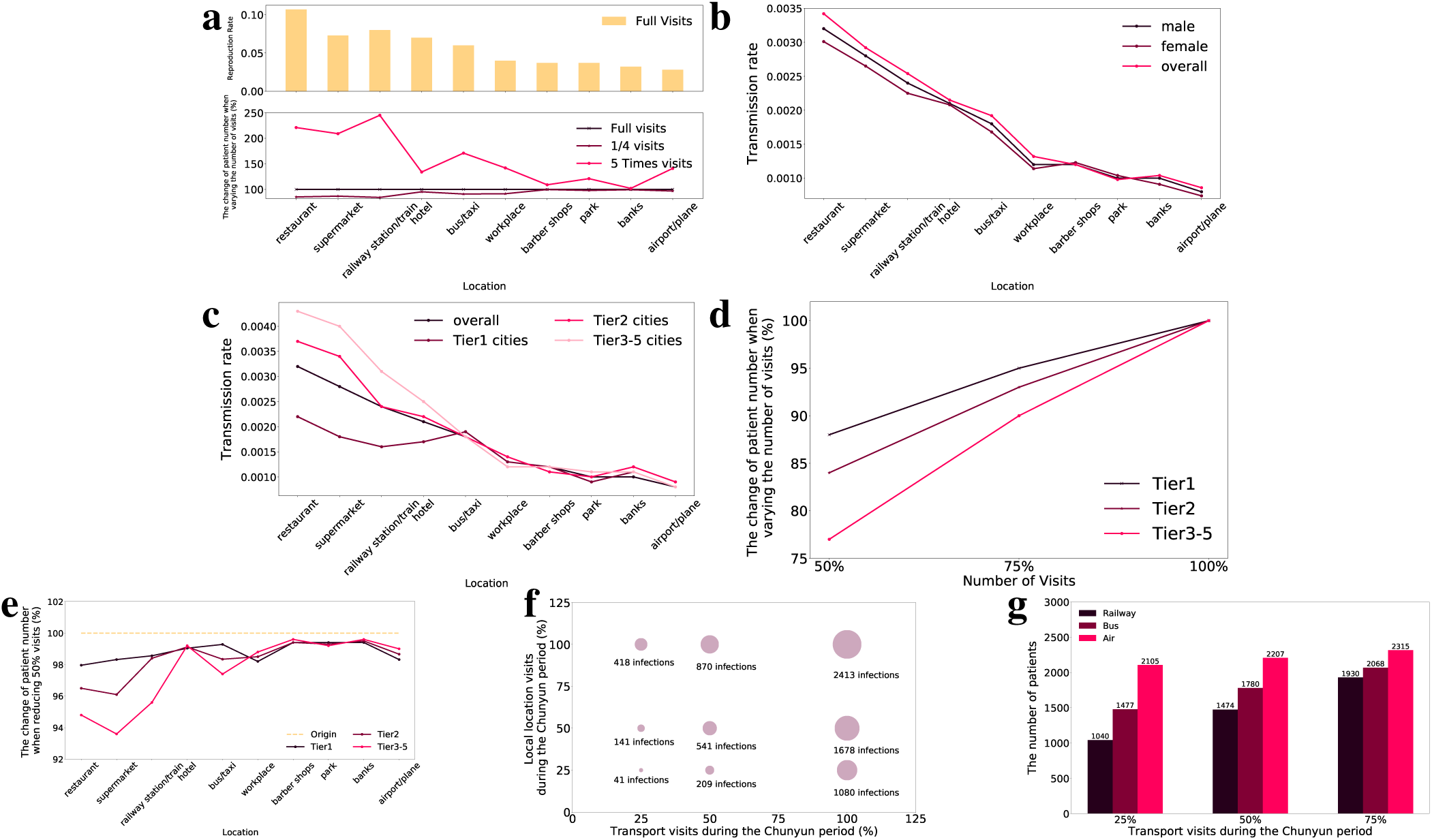
**a**, Transmission rates for different location categories and changes in total infections when varying the numbers of visits to different locations. The highest reproduction number *R* occurs in restaurants (0.11), followed by railway-stations/trains (0.08), supermarkets/grocery-stores (0.073), and hotels (0.071). Reducing visits to railway-stations/train by 75% leads to the greatest total infection decrease (−15.7%), followed by restaurants (−14.8%) and supermarkets (−13.2%). Increasing visits by 5 fold has the greatest impact for railway-stations/train (×2.45), restaurants (×2.21) and supermarkets (×2.09). **b**, Transmission rates of males and females for different locations. The rates are generally higher for males than for females, and parks were the only locations where transmission rates were lower for males than for females. **c**, Transmission rates for different location categories in cities of different tiers. For community locations, tier2 and tier3-5 cities have significantly higher transmission risks than tier1 cities. We postulate that this is because temperature tests and mask-wearing are more strictly imposed in tier1 cities. **d**, Patient number changes when reducing all visits for different city tiers. Reducing visits of all locations in tier1, tier2 and tier3-5 cities with rate *η* = 50% respectively leads to decreases of 12%, 16% and 23% of total infections. **e**, Changes in patient numbers when reducing 50% visits for different locations in different cities. In tier3-5 cities, reducing visits to community locations such as restaurants and supermarkets leads to the greatest decrease in infections, significantly greater than transportation, but for tier1 cities, transportation if of equal importance to community locations. **f**, Reducing visits in community locations and transportations leads to a complementary effect in reducing infections during the Chunyun period. If the increase in transportation is reduced by half and the visits to community locations are increased × 2, the number of infections decreases to 870, compared with original infections 2,431. The number of infections is further decreased to 418 if the increase in transportation is reduced by three quarters *η* = 0.25. With the increase in both community locations and transportations reduced by three quarters during the Chunyun period, there only remain 41 infections. **g**, Restriction on air transport has very limited effects, but restrictions of transport by rail and bus significantly decrease the number of infections.

### Effects of Policies Limiting Visits to Different Location Categories

To further evaluate the effect of mobility increasing and decreasing for different types of locations on the overall number of infections, we conducted simulations on counterfactual networks based on the learned SEIR model. Specifically, we first considered the original network from April 2020 to January 2021. For each category of locations, we constructed counterfactual networks by scaling the magnitude of visits for a single category of locations, but kept visits to other categories of locations stable. The trained SEIR model was then applied to the newly constructed counter-factual network to determine the number of infections. Results are shown in Fig.2a. The number of total infections decreases by 15.7% when reducing visits to railway-stations/trains, followed by restaurants (−14.8%), supermarkets (−13.2%), workplaces (−9.3%), buses/taxis (−9.1%), hotels (−4.7%), airports/airplanes (−3.2%), parks (−2.1%), banks (−0.7%), and barber shops/hairdressers (−0.4%). Generally, reducing visits will reduce the number of infections, which consistent with previous findings [22, 23, 24, 25]. As shown in results, though the transmission rate is lower for railway travel than for community locations such as restaurants and shopping centers, reducing the frequency of travel is effective in a manner similar to that of reducing visits to community locations. This is because traveling facilitates coronavirus transmission across cities, and spreads virus to cities that originally do not have infections. Furthermore, reducing rail travel is substantially more effective than reducing travel by air (−15.7% vs. −3.2%, respectively). We postulate that this is because transmission rate is lower for flight travel than for rail travel, as described above. Moreover, the population from bottom-tier cities comprises a larger proportion of passengers by rail than by flight. As shown below, transmission rates tend to be higher for bottom-tier cities than for top-tier cities. When the magnitude of visits increases by five fold, we observe ×2.45 of total infections for railway-stations/trains, followed by restaurants (×2.21), supermarkets (×2.09), buses/taxis (×1.71), workplaces (×1.42), airports/airplanes (×1.41), hotels (×1.34), parks (×1.21), barber shops/hairdressers (×1.09), and banks (×1.02).

### Economic Tiers of Cities

Based on population size, GDP, and administrative hierarchy, the Chinese city tier system^3^ classifies cities into 5 tiers, with a total number of six categories from tier 1 to 5 with an additional new tier1. We merged tier1 and the new tier1, forming the top-tier category, and tiers 3-5 to form the bottom-tier category, leading to a total number of three tier categories for cities. Transmission rates for different city tiers are shown in Fig.2c. As can be seen, for categories of community locations, tier2 and tier3 cities have significantly higher transmission rates, compared with tier1 cities.

### Effects of Policies of Limiting Mobility of People in Cities of Different Economic Tiers

To examine the influence of mobility restrictions on distinct location categories in cities within different economic tiers, we performed simulations on counterfactual networks based on the learned SEIR model. Results are shown in Fig.2d. Reducing visits of all locations in tier1, tier2 and tier3 cities with rate *η* = 50% respectively leads to decreases of 12%, 16% and 23% in total infections, respectively. Further simulations were conducted with regard to individual location categories in different cities. Fig.2e shows the results. For tier3 cities, reducing visits to community locations (e.g., restaurants and supermarkets) leads to the greatest decrease in the number of infections (significantly greater than transportation). In contrast, for tier1 cities, limitations of transportation (e.g., travel by rail and air) is equally important to limitations of visits to community locations. We postulate that this is because people travel more frequently in tier1 cities than in cities of other tiers. Transportation is therefore as risky as visiting community locations such as restaurants in tier1 cities.

### Policies Regarding Chunyun

Chunyun is a period of travel in China with extremely high traffic load around the Chinese New Year. It usually starts before the New Year’s Day and lasts for approximately 30-40 days. In addition to the marked increase in traffic loads within and across cities, the intensity of local mobility also increases significantly due to social activities such as family get-togethers. We used *baidu immigration*^4^ to obtain the transportation data for previous years. By comparing average traffic load in the Chunyun period with that of the whole year, we found that the greatest spikes occur 3 days before the spring festival, lasting for 3-4 days, and then 4 days after the spring festival lasting for a further 3-5 days. During the spike period, we observe average increases of ×5.2 for rail transport, ×8.9 for road transport, and ×3.8 for air transport.

To simulate the situation with no interventions taken during the Chunyun period, along with examining the effects of different policies, we used the network 1 week before the Spring Festival (February 4, 2021) as the initial state, and then constructed networks regarding different policies. The learned SEIR model was run on the constructed counterfactual networks to examine the outcomes of different policies. As shown in Fig.2f and Fig.2h show the results and observations were as follows: (a) With no interventions taken, where there are increases of ×5.2 for rail transport, ×8.9 for road transport, and ×3.8 for air transport during the spike period, and an increase of ×2 for visits to community locations during the entire Chunyun period, we observe a total number of 2,413 infections during the Chunyun season (February 4, 2021 to March 8, 2022); (b) With the increase in transports reduced by half, and the increase in visits to community locations remaining ×2, the number of infections is decreased to 870; (c) The number of infections is further reduced to 418 if the increase in transport is reduced by three quarters *η* = 0.25; (d) If there is no intervention with traffic load, and we reduce visits to community locations by half, the number of infections remains high (1,678); (e) With no increase in transport, visits to community locations must increase by 9.4 times to reach the number of infections in (d); (f) Restrictions on air transport have limited effects, but restrictions on rail transport significantly decrease the number of infections. Due to the relatively high transport load by buses, restrictions are also important, but less effective than those for rail travel. The explanations are twofold: (1) transportation facilitates transmissions across cities, and spreads virus to cities that originally do not have infections; (2) Because transmission rates in family households are consistently high and are generally unaffected by intervention policies, infections from transports can consistently contribute to the infections through family households even when community locations are locked down.

Therefore, policies that discourage traveling by rail and road are crucial to avoid another wave of breakouts during the Chunyun season. To compensate for the economic loss of traveling during the Chunyun season, restrictions on visits to community locations can be loosened as they contribute substantiallty less to infections, compared with traveling.

### Effective COVID-19 Vaccine Delivery

Due to the supply shortage of vaccine supply, it is important to design effective vaccine delivery strategies to effectively allocate vaccines across population groups e.g., age, sex and city. *V E* denotes vaccine efficacy, which is the proportionate reduction in disease attack rate (AR) between the unvaccinated (ARU) and vaccinated (ARV) groups: *V E* = 1− *ARU/ARV*. In the SEIR model, vaccine interferes with the transitioning stage from susceptible to exposed, where the transmission rate is reduced from *β* to *β*(1 −*V E*). We set the VE value for the general Chinese population to 0.92 [26].

The problem of finding the optimal vaccine delivery strategy can be transformed to an optimization problem, where given a fixed number of vaccines smaller than the population, we search for the optimal vaccine allocation 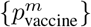 over different population groups *{m}*, leading to the minimum number of infections or deaths. Let 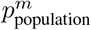 denote the proportion of *m* in the overall population. For category *m*, the proportion of population within *m* receiving the vaccine is 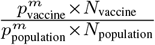. Setting a reduction in the number of deaths as the objective, different fatality rates (probability of dying if infected by the virus) for different age groups should be considered. The fatality rates *f*_*m*_ for children, youths, adults and seniors are respectively set to 0.015%, 0.03%, 1.2% and 11.2%, respectively [27].

### Vaccine Allocation among Different Age Groups

We first explored the distribution of vaccine delivery over different age groups by conducting simulations. Let *α* denote the ratio between the total number of vaccines and the population size. *α* = 0.01 means 1% of the population can be vaccinated. Fig3.a shows that given limited number of vaccines, children and youths should be the last to be vaccinated: as the proportion of children and youths 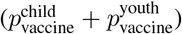 increases, we observe smaller decrease in infections/death. Specifically, if all vaccines are allocated to children and youths, the total number of infections decreases by 0.43% for *α* = 0.01 and 5.1% for *α* = 0.1. For deaths, the decrease are 0.14% for *α* = 0.01, and 2.0% for *α* = 0.1 when all vaccines are allocated to youths and children. Because the fatality rates for children and youths are extremely low, the decrease in deaths is actually caused by the decrease in death of from adults and seniors coming into contact with children and youths. Fig3.b shows results for allocations between adults and seniors. When optimizing for the number of infections, as can be seen, for both *α* = 0.01 and *α* = 0.1, all vaccines should be allocated to adults, achieving an optimal decrease of 2.93% of total infections for *α* = 0.01, and a decrease of 37.3% for *α* = 0.1. This is because adults contribute more to the spread of virus due to their high levels of activity and mobility. When optimizing for the number of deaths, the outcome is different: although adults contribute more to the spread, compared with seniors, they have a significantly lower fatality rate. The best strategy should involve a tradeoff between slowing virus transmission (and thus vaccinating adults) and considering the high fatality rate for seniors (and thus vaccinating seniors). As shown in the figure, when *α* = 0.01, the largest decrease of deaths is obtained when 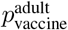and 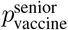 are respectively set to 0.1 and 0,9, respectively, leading to a total decrease of 4.52% in number of deaths. When *α* = 0.1, the largest decrease in deaths is obtained when 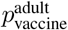 and 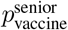 are respectively set to 0.3 and 0.7, leading to a total decrease of 57.2% in the number of deaths.

**Figure 3:**
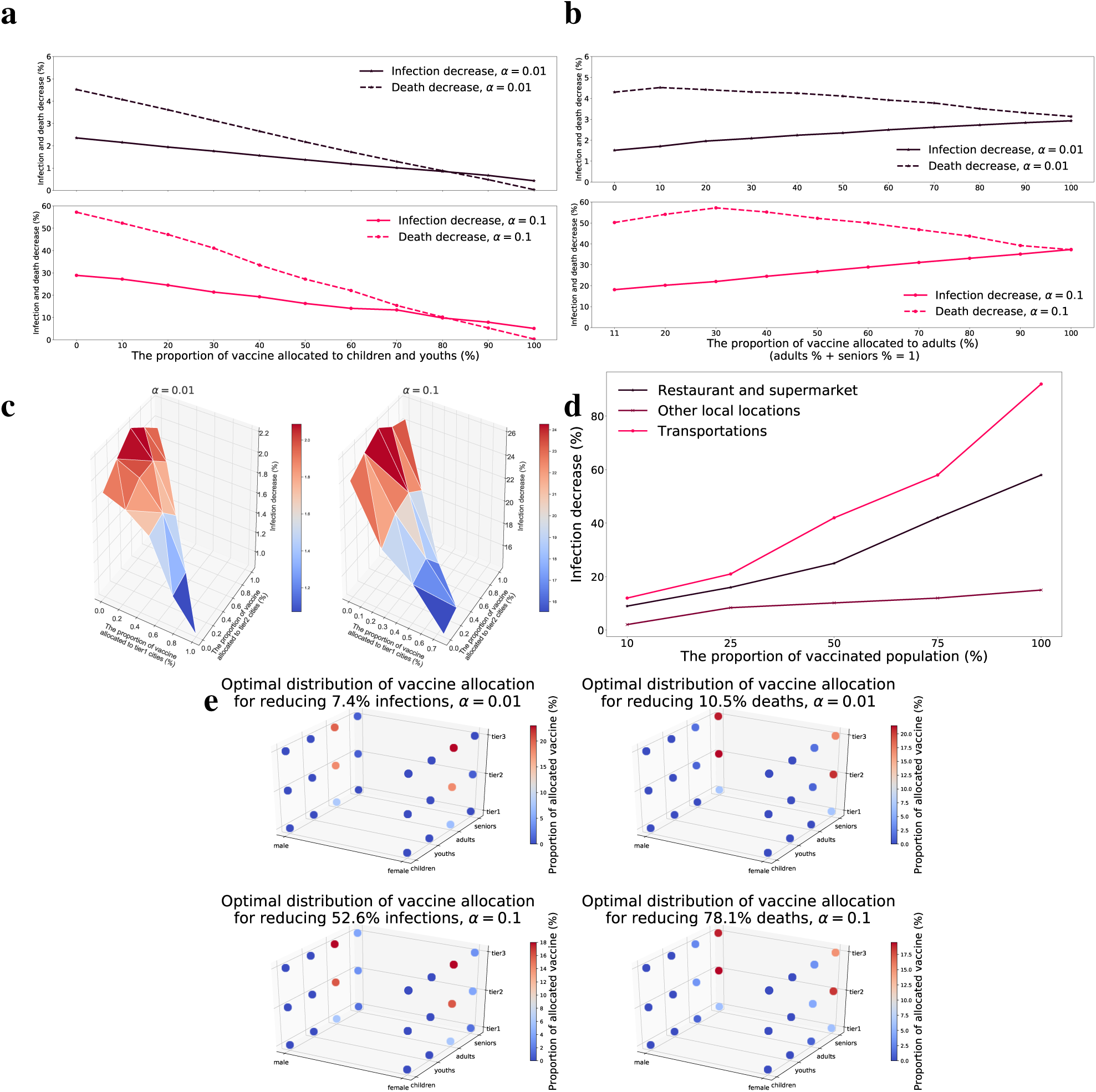
**a**, Given a limited number of vaccines, children and youths should be vaccinated last: as the proportions of vaccinated children and youths increases, we observe lower infection/death decrease. If all vaccines are allocated to children and youths, the infection decrease is 0.43% for *α* = 0.01, and 5.1% for *α* = 0.1. For deaths, the decrease is 0.14% for *α* = 0.01, and 2.0% for *α* = 0.1. **b**, The effects of vaccine allocation between adults and seniors. When optimizing for the number of infections, for both situations where *α* = 0.01 and *α* = 0.1, all vaccines should be allocated to adults, achieving a decrease of 2.93% in total infections for *α* = 0.01, and a decrease of 37.3% for *α* = 0.01. When optimizing for the number of deaths, when *α* = 0.01, the largest decrease of deaths is obtained when 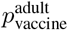 and 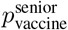 are respectively set to 0.1 and 0.9, leading to a total decrease of 4.52% in the number of deaths. when *α* = 0.1, the largest decrease is obtained when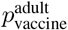 and 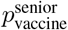 are respectively set to 0.3 and 0.7, leading to a total decrease of 57.2%. **c**, People from cities in the second and third tiers should be the first to receive the vaccine. The greatest infection decrease occurs When 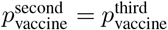 for both *α* = 0.01 and *α* = 0.1.**d**, Vaccinating traveling populations is the most effective means of reducing the proportion of infections. People who travel frequently should therefore be vaccinated first. People who visit restaurants and supermarkets should be vaccinated second. For the remaining community locations, vaccinating visitors is less effective. **e**, Combining age, sex, and economic tiers of cities, when *α* = 0.01%, the optimal combination leads to a decrease of 7.4% in the number infections, where female adults from tier3 cities receive the largest (23%) proportion of vaccines, and to a decrease of 10.5% in the number of deaths, where male seniors from tier3 cities receive the largest (21%) proportion of vaccines. When *α* = 0.10%, the optimal combination leads to a decrease of 52.6% in the number of infections, where female adults from tier3 cities receive the largest (18%) proportion of vaccines, and to a decrease of 78.1% in the number of deaths, where male seniors from tier2 cities receive the largest (19.5%) proportion of vaccines.

### Vaccine Allocation among Cities of Different Economic Tiers

Assuming that the age distribution is identical in cities of different tiers, we searched for the optimal distribution for vaccine delivery over cities of different economic tiers, namely, 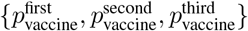. As can be seen from Figure 3c, we find that people from cities in the second and third tiers should be the first to receive the vaccine. We postulate that this is because tier1 cities have a stricter mask-wearing, temperature-testing, and traveler quarantine policies. The greatest decrease in infection occurs when 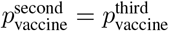.

### Vaccine Allocation between Males and Females

On one hand, males have higher infection and fatality rates, and higher mobility, indicating that males should be vaccinated first. On the other hand, females tend to exhibit a greater immune response that can facilitate vaccine efficacy, indicating that vaccinating females is more effective given limited vaccine availability. Based on our simulation results, where the VE of females is set to 1.2-fold greater than that of males [28] we found that the influence of vaccine distribution according to sex is minimal. For *α* = 0.01, the decreases in total infections are 1.89% and 1.92% when all vaccines are given to males and females, respectively. For *α* = 0.1, the decreases in the number of deaths are 2.34% and 2.29% when all vaccines are given to males and females, respectively.

### Vaccine Allocation based on Location

Because we do not have precise statistics for 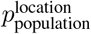, here we only evaluate the impact of vaccinating certain proportions of the population visiting different location categories. The locations are divided into three categories, restaurants+supermarkets, other community locations, and transportation. As shown in Fig.3d, vaccinating the traveling population is the most effective means of reducing the proportion of general infections. People who travel frequently should therefore be vaccinated first. People who visit restaurants and supermarkets should be vaccinated second. People who visit restaurants and supermarkets should be vaccinated second. For the remaining community locations, vaccinating visitors is less effective.

### Combining Age, sex and Economic Tiers of Cities

Fig.3e shows the distributions of vaccines allocated to different population groups considering age, sex and city tiers to achieve the minimum numbers of infections and deaths. When *α* = 0.01%, the optimal combination leads to a decrease of 7.4% in infections, where female adults from tier3 cities receive the largest (23%) proportion of vaccines, and to a decrease of 10.5% in deaths, where male seniors from tier3 cities receive the largest (21%) proportion of vaccines. When *α* = 0.10%, the optimal combination leads to a decrease of 52.6% infections, where female adults from tier3 cities receive the largest (18%) proportion of vaccines, and to a decrease of 78.1% deaths, where male seniors from tier2 cities receive the largest (19.5%) proportion of vaccines.

## Discussion

This study has several limitations. First, the analyzed dataset has selection bias. Available action tracking reports are all from the time period after April 2021 when there were only sporadic breakouts in China. Reports during the first wave of outbreaks in January and February 2021 are not available, when there are substantially large numbers of confirmed cases, are not available. Therefore, the results of this study are limited with regard to both location and time. Second, because we were only able to collect reports for only a proportion of the confirmed patients, important aspects might have been missed: for example, since reports for international travelers were not available and were therefore excluded from the dataset. Thus, the observation that flight is a relatively safe means of transportation is limited to domestic flights and cannot be extended to international flights. Third, because all data were collected after April 2021 in China, the results and conclusions of this study are limited to the situation where the pandemic is generally well-contained, and mask-wearing and temperature testingare generally adopted. Therefore, the results cannot be readily generalized to situations with high rates of new daily cases.

Despite these limitations, this work presents the first attempt to analyze the detailed action reports of confirmed cases with comprehensive tracking information, and captures important aspects of transmission of SARS-CoV-2, such as community location, sex, age and city tiers. Capturing these aspects leads to high predictive accuracy, which demonstrates the superiority of the approach. Our results provide insights for control and contact tracing policies. For example, our results suggest that policies regarding limitations on visits to different location categories should be different, and that more limitations should be placed on locations where the mask-wearing cannot be strictly enforced such as restaurants. Regarding contact tracing policies, our results suggest that more emphasis should be placed on tracing people who visit restaurants with confirmed cases. Our results also provide suggestions on policies regarding different demographic groups and cities. For example, for daily essential visits, such as those involving grocery stores, policies should encourage visits by children and youths, rather than by adults and seniors. More importantly, our research provides a statistical foundation for vaccine delivery under conditions of vaccine shortage. Our work will be helpful in designing effective policies regarding interventions, reopening, contact tracing and vaccine delivery in the “new normal” world following COVID-19 spread.

## 2 Methods

### 2.1 Datasets

Action tracking reports are publicly available, and were originally published to the public by provincial Centers for Disease Control and Prevention to warn local residents of locations that confirmed patients had been to, and were broadcast by newspapers and on online social media. ^5^ Patients’ names are anonymized in the reports.

We collected action tracking reports from the websites of provincial Centers for Disease Control and Prevention. We first obtained all contents posted on the websites of 31 (all except Taiwan) provincial Centers for Disease Control and Prevention in China since Jan 2020, as shown in Tab. 1. Posts with titles containing the keyword “轨迹” are retrained, with the rest contents discarded. The remaining posts were further manually examined, and texts chunks corresponding to tracking details were selected. We are able to identify tracking reports for 1,752 patients. The anonymized data are then transformed to obtain population-level statistics, and the research is not based on information at the individual level.

**Table 1:**
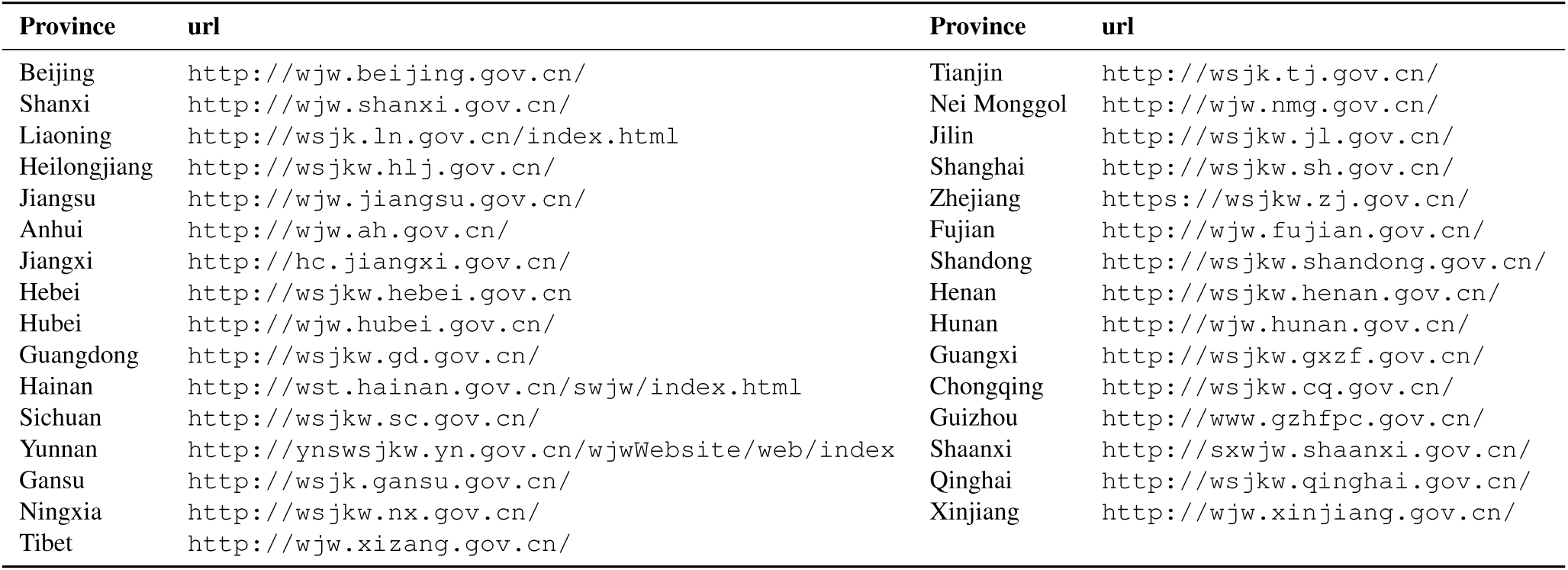
Website URLs of 31 (all except Taiwan) provincial Centers for Disease Control and Prevention in China.

### 2.2 NLP Tools to Transform Unstructured Action Reports to Structured Networks

We use NLP models to extract locations, time and relations between them. We manually labeled 10 percent of the reports by identifying containing locations, times, and whether a specific time corresponds to the patient visits a specific location. Based on labeled data, we train NLP models to extract locations, time and relations from the unlabeled data. We use BERT [29] as the model backbone. Extracted entities and relations will be further verified by human annotators. This process saves the efforts to label the entire corpus.

#### Model training

The input text sequence **x** = *{x*_1_, *…, x*_*n*_*}* contains various entities such as time and locations. For example, if the input sequence is **x** =*{* On, January, 2, she, did, not, go, out*}*, then “January 2” is an entity of type *time*, “she” is an entity of type *person*. These entities should be extracted from the input text sequence. To this end, we adopt the `BIEOS` scheme to predict the label for each of the input units. B, I, E, O and S respectively stand for “Beginning”, “Intermediate”, “End”, “Outside” and “Single”, and they are combined with each of the pre-defined entity types. Take the *time* type as an example. All the labels regarding *time* are `B-TIME`, `I-TIME`, `E-TIME`, `O` and `S-TIME`. If there are *m* different entity types, there will be a total number of *k* = 4*m* + 1 labels, forming the label set 𝒴. The model assigns one label from 𝒴 to each input unit *x*_*i*_ based on whether it is part of an entity and which entity it belongs to. In the example of **x** = *{*On, January, 2, she, did, not, go, out *}*, the ground-truth label sequence is *{ŷ*_1_, *ŷ*_2_, *ŷ*_3_, *ŷ*_4_, *ŷ*_5_, *ŷ*_6_, *ŷ*_7_, *ŷ*_8_*}* = *{*`O`, `B-TIME`, `E-TIME`, `S-PERSON`, `O`, `O`, `O`, `O` *}*, which means “January” is the beginning of a *time* entity and “2” is the end of the *time* entity, along with “she” being a single *person* entity. We encode the ground-truth label *ŷ*_*i*_ into a one-hot vector **ŷ**_*i*_ of length *k*, where the dimension for the corresponding label is 1 and other dimensions are 0. With the ground-truth one-hot label vector **ŷ**_*i*_ for each input text unit and the label distribution vector **y**_*i*_, we can train the model using the cross entropy loss:

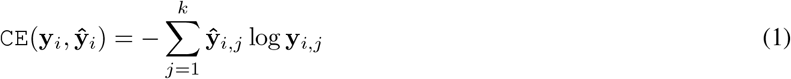

We use Adam[30] to optimize the loss. Besides entities, we also consider location-time relations, i.e., whether a location entity *e*_*l*_ is associated with a time entity *e*_*t*_, signifying that the person visited location *l* at time *t*. To this end, we first represent each extracted entity *e* by concatenating the high-dimensional vector representation of its head unit **h**_*e*.*h*_ and that of its tail unit **h**_*e*.*t*_, and then we transform the result using a learnable matrix **W**_*r*_, which gives the final entity-specific vector representation **r**_*e*_:

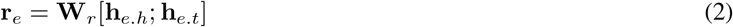

To determine the relation between a location entities *e*_*l*_ and a time entity *e*_*t*_, we apply dot-product to their entity-specific vector representations 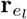 and 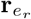 followed by the `sigmoid` function to obtain the probability that *e*_*l*_ and *e*_*t*_ should be associated:

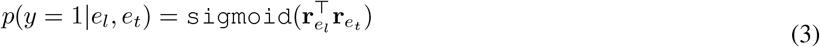

The ground-truth label *y* for a location-time pair is either 0 (no association) or 1 (association), and therefore the model can be trained using the binary cross entropy loss:

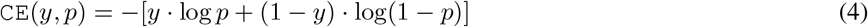

Note that during the location-time decision process, only the learnbale matrix **W**_*r*_ is trained and the base entity extraction model is fixed after trained with Eq.1.

### 2.3 The SEIR Model

#### 2.3.1 Learning

To model the spread of SARS-CoV-2, we use a standard SEIR model with susceptible (S), exposed (E), infectious (I) and recovered (R) states. We follow the standard paradigm for the SEIR model, where a susceptible case is transformed to an exposed case through their contact at location *C* with the transmission rate *β*_*C*_. The exposed state denotes the incubation period during which individuals are infected but are not yet infectious. An exposed case transitions to an infectious case at a rate proportional to the inverse of mean incubation period. Once an individual enters the infectious state, it can spread the virus to susceptible cases. An infectious case transitions to a recovered case at a rate proportional to the inverse of mean infectious period. Once the transition happens, the individual can not get infected or infect others.

For a specific location *c* of category *C*, we use the group of people that visits location *c* at time *t*_visit_ as the basic unit for modeling and trace their actions and disease status. This group of people is denoted by 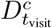. The size of 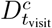 is denoted by 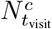. Since we postulate that virus transmission only happens at locations confirmed patients have been to, we only need to model people groups ^*t*^. Among the 1,752 confirmed patients, we collected 7.5k location-time pairs. Since infections are generally sparse, we further postulate that, for group 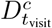, all transitions from S to E in the group happen at location *c* at time *t*. This means that for a susceptible person in 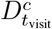, if he does not transition to the exposed state at location *c* at time *t*, he will not get infected in the future.

Since the time each patient being diagnosed is included in and can be extracted from the action tracking reports, we can obtain the number of newly diagnosed (confirmed) cases that have visited location *c* at time *t*, denoted by 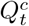. It is worth noting that 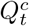 is not the same as 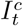, where 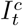 is the number of infections reported at time *t* that have been to location *c*. This is because a patient cannot be diagnosed right after it becomes infectious. We use *I*^*c*^ to denote the number of total infections that have been to location *c*, to bridge the gap between 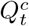 and 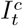:

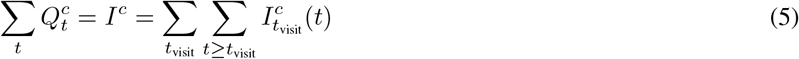

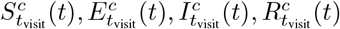 respectively denote the number of susceptible (S), exposed (E), infectious (I) and recovered (R) cases at time *t ≥ t*_visit_ within group 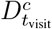. Based on the assumption that virus transmission for 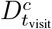 only happens at time *t* at location *c*, transitions from susceptible cases to exposed cases only happen at time *t*_visit_. For time *t* = *t*_visit_, we have:

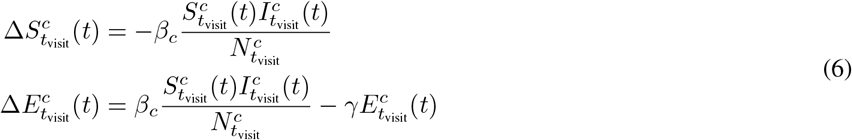

Since we have made the assumption that infections only take place at time *t*, we thus have 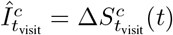.

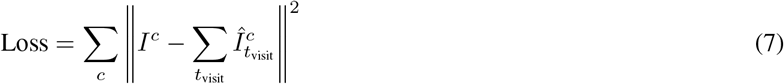

To fit the number of confirmed cases visiting location *c*, we use grid search to search the combination of parameters *β*_*c*_. The optimal value of *β*_*c*_ is obtained with the smallest *L*_2_ loss between the number of reported cases *I*^*c*^ and the sum of predicted infections 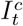:

##### Baseline

A straightforward baseline is to ignore location-level statistics and demographic factors provided by action reports, where predictions on city-level confirmed cases are made only based on mobility data. The model is trained to minimize the *L*_2_ distance between the number of city-level confirmed cases and the predicted number:

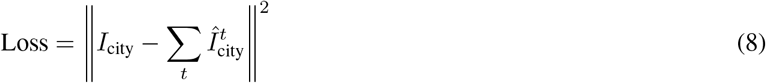

In this way, the system degenerates into the system similar to [20], where only mobility data is used for modeling. As shown in Fig.1d, the baseline model significantly underperforms the proposed model that is based on location-level statistics and demographic factors in terms of prediction accuracy.

##### Validation

For model validation, we first split the time period from March 2020 to January 2021 into consecutive time snippets, with the size of stride set to a week. Each of the snippets lasts a month. We divide all snippets to 80%/20% for training and test, where we train the model based on the 80% snippets, and evaluate the predictive accuracy on the held-out test snippets. As shown in Fig.1d, the used model fits held-out data pretty well, significantly outperforming baseline models.

#### 2.3.2 Age-focused SEIR Model

Let *m* denote the index of the age group, which tasks a value from the age group set Age = *{*child, youth, adult, senior *}*. 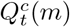 denotes the number of newly confirmed cases at time *t* belonging to age group *m* that have been to location *c*. 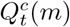 can be readily computed based on action reports. Let *I*^*c*^(*m*) denote the number of infections belonging to age group *m* that have been to location *c*. We have:

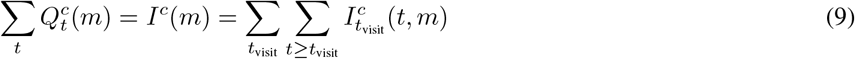

For each location *c*, transmission rates for different age groups are different, denoted by 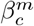, where *m* denotes the index of an age group. 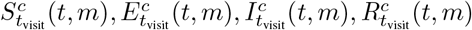 respectively denote the number of susceptible (S), exposed (E), infectious (I) and recovered (R) cases at time *t* ≥ *t*_visit_ within group 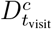 belonging to age group *m*. The age-focused SEIR model is given as follows. For time *t* = *t*_visit_, we have:

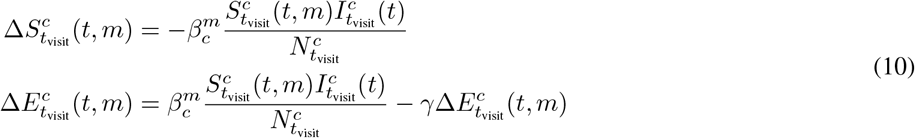

Again, since we have made the assumption that infections only take place at time *t*, we have 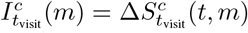. Here we assume that the incubation period and the infectious period for all ages are the same, Similar to the previous section, we use grid search to search the combination of parameters 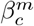 to fit the number of confirmed cases that have visited location *c* of age group *m*:

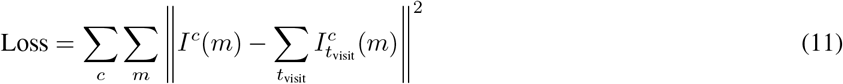

### 2.4 Simulations

Given learned parameters *β* for different locations, city tiers and age groups, we can perform simulations based on graphs. Our simulations are performed on graphs with to 100 million nodes of individuals to best simulate the scenario in China. Nodes of individuals are first clustered into cities of three tiers. Individuals in the same city are connected through time-varying edges indicating visiting the same community location at a specific time, and covid is spread with location-specific and city-specific transmission rates in different locations and cities. Individuals in different cities are connected through time-varying edges indicating transportations, and covid is spread with transportation-specific transmission rates. Simulations were performed on platforms of high performance computing with thousands of cores and terabytes of RAM. During simulations, we use the SEIR model to simulate the status of each individual node.

For each individual node *s*, let state(*s*) denote its corresponding state, taking the value from *S, E, I, R*, which respectively corresponds to the susceptible, exposed, infectious and removed state. For a susceptible node *s*, if it visits location *C* at the time *t*, the probability of transitioning to stage *E* is given by:

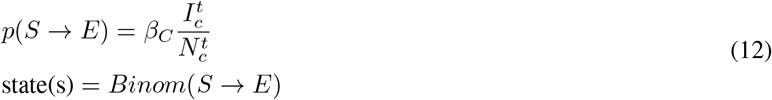

where 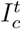 and 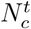 respectively denote the number of infectious nodes and the total number of nodes visiting *C*. If city-tiers and age are consider, city-tier specific and age group specific *β* will be used. For simplification, we use time-consistent 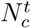 for different locations, which are obtained by averaging the number of to locations belonging to the same location type based on the SmartStep data. For exposed and infectious nodes, the probabilities of transitioning to infectious and removed states are given as follows:

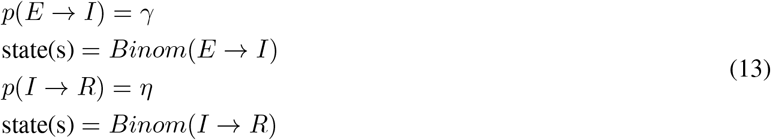

*γ* and *η* are the inverse of average incubation period and infectious period, which are respectively 96h and 84h [10].

#### R0 Estimation

Let *R*_0_ denote the reproduction number for the whole population, and *R*_*C*_ denotes the average number of secondary patients in location category *C* caused by a patient. *R*_*C*_ can be estimated directly from the dataset. Let *H*_*C*_ the total number of patients that have been to category location *C*. For a patient *p ∈ H*_*C*_, let 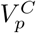 denote the collection of visits of patient *p* to locations of type *C*. For each visit 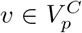, let *N* (*v*) denote the number of confirmed patients occurring at the same location and at the same time. *R*_*C*_ is computed as follows:

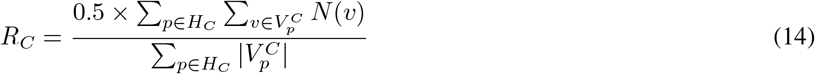

*R*_0_ is computed by summing *R*_*C*_ with weight *F*_*C*_:

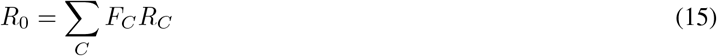

where *F*_*c*_ denotes the average number of a patient visiting location category *C*. We can directly estimate *R*_*c*_ and *F*_*c*_ from the action report dataset. The value of *R*_0_ in this work is 0.37.

For age group, let *R*_*m*_ denote the reproduction rate for age category *m*, which is the average number of secondary patients belonging to age group *m* caused by a patient. *R*_*m*_ for children, youths, adults and seniors are respectively 0.012, 0.028, 0.21, 0.12. *R*_*m*_ is highly correlated with the total population of each age category.

### 2.5 Simulations For Vaccine Delivery

Let *N*_vaccine_ and *N*_population_ denote the number of available vaccine doses and the size of population. Here we make a simplification that each person only needs one dose. Let *M* denote the set of categories on which vaccines are distributed. It can be sex, age or city tiers. Take age as an example, *M* = *{*child, youth, adult, senior*}. m* takes one of the values from 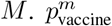 denotes the distribution of vaccine, where the number of children, youths, adults and seniors got vaccinated is *p*^child^ × *N*_vaccine_, *p*^youth^ × *N*_vaccine_, *p*^adult^ × *N*_vaccine_, and 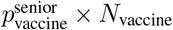. Let 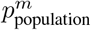 denote the proportion of *m* in the whole population. For category *m*, the proportion of population within *m* getting vaccinated is thus 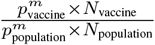. Considering that the value of *V E* can be different for different age categories, the age-specific transmission rate *β*_*m*_ is reduced to *β*_*m*_(1 −*V E*_*m*_), where *V E*_*m*_ denotes the age specific vaccine efficacy. The task of identifying the best vaccine delivery strategy is transformed to an optimization problem. If the objective is minimizing the number of infections, the problem is formalized as follows:

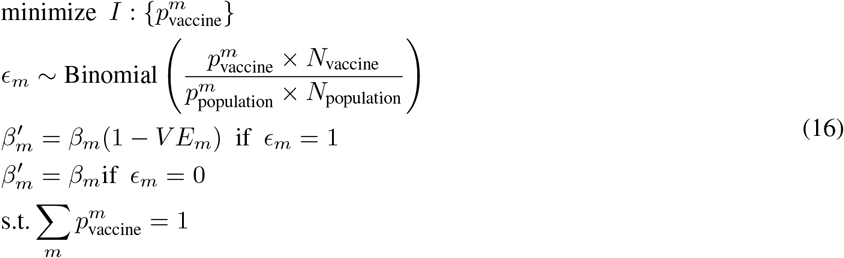

If the objective is minimizing the number of death, age-specific death rates should be considered. The problem is formalized as follows:

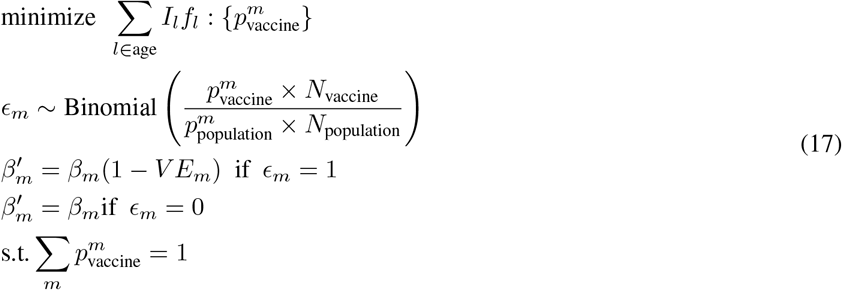

To identify the optimal values of 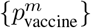, we perform simulations using grid search to obtain the best set of 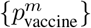 for the number of infections and deaths.

## Data Availability

All the action reports used in this work are publicly available on the websites of provincial centers for Disease Control and Prevention (e.g., http://wsjkw.hlj.gov.cn/ and http://wsjkw.hebei.gov.cn/).
Reports are also reproduced online by news websites
and can be found on social media websites such as Weibo. There are no restrictions to date on the use of the data for research purposes. The SmartStep data are from a commercial product, owned by the third party, and will be available from the corresponding author upon reasonable request.

http://wsjkw.hebei.gov.cn

## 3 Data availability

All the action reports used in this work are publicly available on the websites of provincial centers for Disease Control and Prevention (as listed in the Dataset subsection). Code for accessing the contents of corresponding websites and context filtering will be provided upon publication. The final action tracking report dataset will be released upon publication. The SmartStep data are from a commercial product, owned by the third party, and will be available from the corresponding author upon reasonable request.

## 4 Code availability

Data analysis was performed using Python and Lua. Code is available at https://github.com/ShannonAI/action_tracking_report.

## Footnotes

1 Based on regulations http://www.xinhuanet.com/politics/2021-01/24/c_1127019082.html, reports contain only the tracking information of patients. Personal information such as names, sexes, ages and addresses that makes patients identifiable is not allowed to be disclosed in the reports.

2 Another justification for this assumption is that action reports are collected by city. If one report of a city is found in the collection, it is very likely that reports for all patients in that city are included.

3 https://en.wikipedia.org/wiki/Chinese_city_tier_system

4 http://qianxi.baidu.com/

5 Examples: action tracking reports for diagnosed patients on Jan.19 2021 in the Heilongjiang province, found on the website of Heiongjiang CDC: http://wsjkw.hlj.gov.cn/pages/601a043b4ed1dc8e06c86a10; action tracking reports for diagnosed patients on Jan 5, 2021 in the Hebei province, found on the website of Heibe CDC: http://wsjkw.hebei.gov.cn/syyctplj/375192.jhtml.

